# 3D phenotyping in a Colombian population reveals unique population and ontogenic facial patterns in genetic and rare disorders

**DOI:** 10.1101/2025.02.11.25322000

**Authors:** Mireia Andreu-Montoriol, Marina Pujol, Luis Miguel Echeverry-Quiceno, Estephania Candelo, Álvaro Heredia-Lidón, Eidith Gómez, Ona Roure-Ramis, Max Rubert-Tayà, Xavier Sevillano, Maria Esther Esteban, Aroa Casado, Harry Pachajoa, Neus Martínez-Abadías

## Abstract

**Background:** Approximately 30-40% of genetic and rare disorders manifest with distinct facial patterns. Traditionally, clinical geneticists have qualitatively assessed facial morphology to support preliminary diagnosis and guide confirmatory genetic testing. However, enhancing early diagnostic accuracy through facial biomarkers demands advanced 3D technologies for analyzing facial dysmorphologies, a deeper understanding of condition-specific disruptions in facial development, and a broader inclusion of diverse human populations.

To bridge this gap, we analyzed the 3D phenotypes associated with four genetic syndromes in an admixed Latin American population from Colombia. The sample comprised 47 individuals diagnosed with Down (DS), Morquio (MS), Noonan (NS), and Neurofibromatosis type 1 (NF1) syndromes along with 49 controls within the same age range. For each participant, we generated a 3D facial model using a multi-camera photogrammetric system and registered the 3D coordinates of 21 anatomical facial landmarks. Geometric morphometrics methods were employed to characterize syndrome-specific 3D facial dysmorphologies and to assess their variation compared to controls. We also examined whether these syndromes alter the ontogenetic trajectory of facial growth.

**Results:** Facial shape differed significantly in all syndromes except NF1. Consistently with previous 2D studies, we identified population-specific facial features in Colombian patients that are not reported in individuals of European descent. Pooled 3D analyses revealed a continuous spectrum of facial dysmorphology, with MS displaying the most distinct morphology and an altered ontogenic pattern. Facial size, sex and age were all significant factors modulating facial shape, with diagnosis explaining 14% of variation in facial morphology.

**Conclusions:** Overall, these findings highlight the importance of accounting for interpopulation, sex and ontogenic variation in facial phenotypes to improve the diagnostic utility and of facial biomarkers. Such approaches may contribute to shortening the diagnostic odyssey for individuals with syndromic and rare genetic conditions.

## BACKGROUND

Genetic and rare diseases affect approximately 7% of the global population, corresponding to 500 million people worldwide (1,2). Although each rare disease occurs in a small number of individuals, fewer than 1 in 2,000 individuals, more than 7,000 distinct rare conditions have been identified, each characterized by unique clinical features (3). Early diagnosis is critical for effective genetic counseling, timely intervention, and prevention (4). However, significant diagnostic challenges persist due to limited understanding of underlying genetic mechanisms, overlapping clinical manifestations, and the scarcity of reliable diagnostic tools, particularly during early life (4). To overcome these challenges, researchers are exploring complementary strategies beyond traditional clinical and molecular diagnosis (5). Among these, facial dysmorphology analysis has emerged as a promising approach, combining the analysis of facial imaging algorithms with genomic data to establish a link between patients’ rare variants and potential facial gestalt (6,7).

Approximately 30-40% of rare genetic disorders exhibit distinct facial dysmorphology patterns, often resulting from disruptions in genetic signaling pathways that regulate craniofacial development, such as Sonic Hedgehog (SHH), Fibroblast Growth Factor (FGF), Bone Morphogenetic Protein (BMP), Wingless-related integration (Wnt), and Transforming Growth Factor β (TGF β) (8–15). These facial traits can range from subtle to severe (16), and previous research demonstrates that they provide valuable diagnostic cues (12–14).

Traditionally, facial assessment has relied on qualitative description and basic anthropometric measurements to guide preliminar diagnosis and confirmatory genetic testing (17–20). However, the variability and complexity of facial features both across and within syndromes demand highly trained specialists and remain challenging to evaluate (20). Recent advances in computer vision, machine learning, and geometric morphometrics now allow for more objective and automated facial analysis (12–14,21–27). Tools such as Face2Gene have shown significant potential and are increasingly integrated into clinical workflows (12–14). Nevertheless, current tools still face limitations in capturing the 3D complexity, evolution, and diversity of facial phenotypes (13,28).

First, most automated methods use two-dimensional (2D) images, which are easier to collect but fail to capture depth-related facial traits that are often altered in genetic disorders (12). In contrast, three-dimensional (3D) methods, though less accessible, can more effectively characterize disorder-specific facial patterns (13). Recent studies using 3D geometric morphometrics have revealed detailed phenotypic signatures in rare and genetic disorders, as well as differences between these patients and control individuals (13,27). Moreover, subtle yet significant shape differences have also been detected in patients with psychotic disorders (29).

Second, although facial dysmorphologies often emerge during embryonic development and evolve throughout life (30), the influence of growth on facial shape in rare disorders has been minimally explored. Understanding whether these conditions drive age-related growth deviations is crucial for further adjusting diagnostic and prognostic models (11).

And third, the limited availability of large-scale datasets for training diagnostic algorithms has resulted in an overrepresentation of individuals of European descent (13,31,32), leaving African, Asian, and Latin American populations underrepresented. This imbalance is also evident in key medical references, such as Smith’s Recognizable Patterns of Human Malformation, which largely omits Latin American individuals (16). Consequently, diagnostic algorithms based on European-centered data fail to capture the full genetic and phenotypic diversity of other populations. Increasing evidence suggests that incorporating diverse ancestries into biomedical research enhances the identification of genetic variants associated to anthropometric traits (33), craniofacial morphology (34) and disease susceptibility (35), ultimately improving diagnostic accuracy.

### 3D facial shape variation across syndromes and ontogenesis in an admixed population

Previously, we analyzed 2D facial dysmorphologies associated with genetic and rare disorders in a highly admixed Colombian cohort of pediatric and adult individuals (7). That study revealed population-specific traits and demonstrated that current diagnostic algorithms based on 2D facial morphology performed less accurately in this cohort than in European populations (7). Building on those findings, we now apply 3D analyses to re-examine facial phenotypes of Down, Morquio, Noonan, and Neurofibromatosis type 1 syndromes within the same population.

Down syndrome (DS, OMIM 190685) is a genetic condition caused by trisomy of chromosome 21 (36), occurring in approximately 1 in 800 births worldwide (37). This chromosome imbalance leads to intellectual disability and multi-systemic complications, including cardiovascular, respiratory, digestive, neurological, and musculoskeletal issues (38). The craniofacial phenotype is characterized by brachycephaly and midfacial flattening (39–41), along with oblique palpebral fissures, hypertelorism, a broad nose with a depressed bridge, a small open mouth, macroglossia, micrognathia, and a narrow palate, all of which can impair breathing, feeding, and speech (38,40,41).

Morquio syndrome (MS, OMIM 253000), or mucopolysaccharidosis type IVA, is an autosomal recessive disorder with a global birth prevalence of approximately 1 in 200,000-300,000 (42). Colombia reports one of the highest prevalence rates worldwide (0.68 per 100,000 live births), likely due to founder effects (43,44). MS is caused by mutations in *GALNS*, a gene encoding an enzyme required for glycosaminoglycan metabolism (42). Clinical features include short stature, skeletal deformities, joint hyperlaxity, and coarse facial features such as frontal bossing, flat nasal bridge, thick lips, macroglossia, corneal opacity, glaucoma, and hypertelorism (42,45).

Noonan syndrome (NS, OMIM 163950) and Neurofibromatosis type 1 (NF1, OMIM 162200) are RASopathies (46), caused by alterations in the regulation of the RAS-MAPK pathway (47). NS is an autosomal dominant condition (1 in 1,000-2,500 live births) (47) caused by mutations in the *PTPN11* (50%), *SOS1* (23%), *RAF1* (5%), or *KRAS* (<5%) genes (48). Clinical features include mild intellectual disability, cardiac defects, skin pigmentation, and skeletal anomalies (49). Distinct facial traits include a wide forehead, hypertelorism, ptosis, epicanthic folds, arched eyebrows, a broad neck, low posterior hairline, and low-set posteriorly rotated ears (50,51).

NF1 is an autosomal dominant disorder associated with a prevalence of 1 in 4,000 live births (52). It is caused by loss of function in *NF1*, a tumor suppressor gene regulating RAS proteins (52). Clinical features include café-au-lait spots, axillary freckling, Lisch nodules, optic gliomas, and skeletal anomalies (52,53). Craniofacial features include macrocephaly, proptosis due to sphenoid wing agenesis or optic pathway gliomas, maxillomandibular deformities, and dental anomalies (54).

Clinical manifestations associated with these genetic and rare disorders are known to vary across population ancestry (7,55–57). The objective of this study is to quantify 3D facial dysmorphologies associated with DS, MS, NS and NF1 in an admixed Colombian population. Using 3D geometric morphometrics, we compared patients and control individuals to define disorder-specific and ontogenetic facial patterns. We hypothesize that the characterization of these features in an admixed population could improve early, accurate, and unbiased diagnosis, supporting more personalized clinical management.

## METHODS

### Population sample

The study sample, collected in 2021 in Cali, Colombia (7) comprised 96 individuals, including children and adults (from 2 to 51 years) of both sexes (Table 1). Controls were recruited at Universidad ICESI and Colegio Ecológico Scout; whereas individuals diagnosed with Down, Morquio, Noonan and Neurofibromatosis type 1 syndromes were recruited through the clinical genetics service at Hospital-Fundación Valle del Lili, a tertiary referral center for genetic and rare disorders. Inclusion criteria for patients required that clinical diagnoses were confirmed by molecular genetic testing. Clinical history of craniofacial trauma and/or surgery were exclusion criteria for all participants.

**Table 1:** Cohort composition by diagnostic groups and sex.

| Syndrome | Female | Male | Total |
| --- | --- | --- | --- |
| Down | 11 | 8 | 19 |
| **Morquio** | 5 | 3 | 8 |
| **Noonan** | 5 | 3 | 8 |
| **Neurofibromatosis type**<br>**1** | 6 | 6 | 12 |
| **PATIENTS** | 27 | 20 | 47 |
| **CONTROL** | 34 | 15 | 49 |
| **TOTAL** | 61 | 35 | **96** |

The study was approved by the Ethics Committee for Human Research at Universidad Icesi (approval record no. 309), the Bioethics Committee of the Universitat de Barcelona (IRB03099-CER032514) and adhered to national and European research guidelines, following the standards for medical research in humans recommended by the Declaration of Helsinki. All participants provided written informed consents before enrollment. For minors, parental or legal guardian consent was obtained for the collection of facial photographs and clinical data.

### Acquisition of Facial Images and Anatomical landmarks

Facial data were acquired via photogrammetry, a non-invasive, efficient, and cost-effective technique for generating high-resolution 3D models (13,41). The setup consisted of 10 synchronized digital cameras (SONY Alpha 58 +18-55) mounted on five tripods at two heights, arranged in a semicircle one meter from the subject (41). Cameras were triggered simultaneously via remote control to capture images of the participants from multiple angles. People were asked to maintain an upright position with a neutral facial expression. Overlapping images were processed in AgiSoft PhotoScan to generate textured 3D surface meshes, which were scaled and saved as Polygon File Format (PLY) files.

To analyze the facial morphology associated with DS, MS, NS and NF1, 21 anatomical facial landmarks were manually placed on all individuals using 3DSlicer 5.0.2 (Figure 1; Additional file 1) (58). A single trained observer conducted repeated measurements on a subsample of 25 individuals (five per group) across five sessions at 48-hour intervals. Intra-observer error was significantly lower than biological variation (p-value <0.010), with a mean error <2 mm, within the accepted threshold for craniometric studies (59). Landmark coordinates were subsequently used for morphometric analyses.

**Figure 1.**
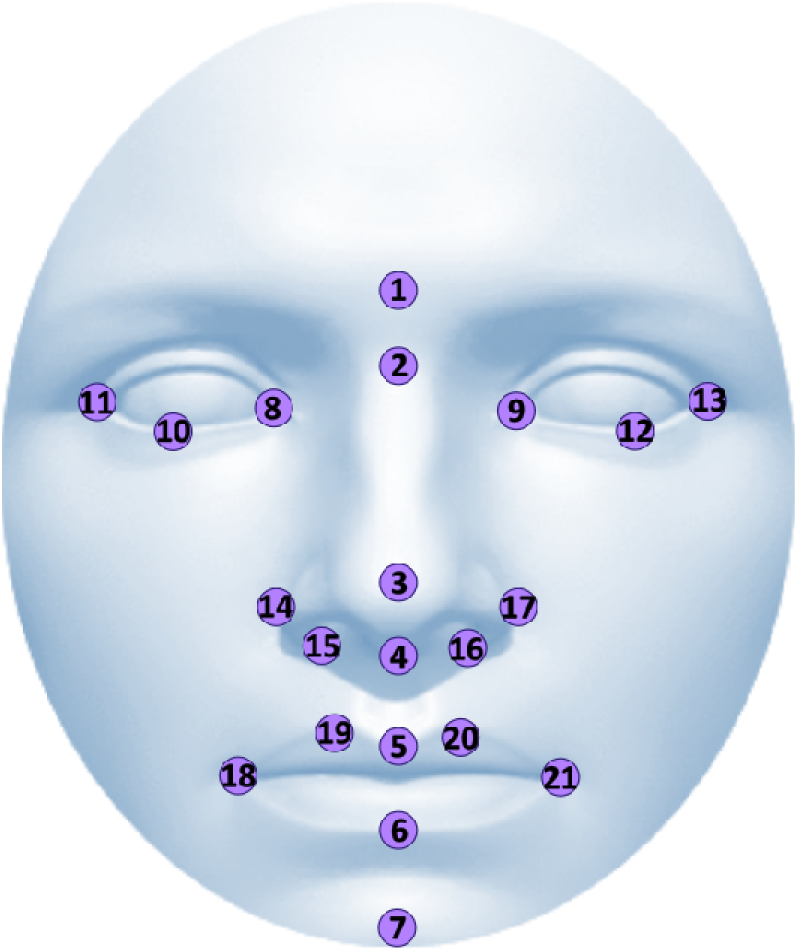
3D facial reconstruction showing the 21 anatomical landmarks (numbered symbols). Landmark definitions are provided in Additional file 1.

### Quantitative Facial Shape Analyses

Facial shape differences were first evaluated within each syndrome, by comparing patients and controls. Generalized Procrustes Analysis (GPA) was applied to the 3D landmark coordinates (60), standardizing configuration through translation, scaling, and rotation to achieve optimal superimposition (60). The resulting Procrustes coordinates were analyzed via Principal Component Analysis (PCA). Morphological variation was visualized in the morphospace defined by the first two principal components (PCs). Given the limited sample size, analyses were performed without separating by sex to preserve statistical power. Nevertheless, sex was visually distinguished in PCA plots and incorporated as a covariate in subsequent analyses. Overlapping patient-control groups indicated minimal dysmorphology, whereas distant clusters signified pronounced phenotypic differences. Procrustes distances between the mean shape of patient-control groups were calculated as the square root of the summed squared differences between homologous landmarks (61). Statistical significance was assessed via permutation tests (1,000 iterations).

Subsequently, GPA and PCA were performed on the pooled dataset to explore facial shape differences across syndromes. A Procrustes ANOVA using the procD.lm function in the geomorph R package, assessed the influence of diagnosis, size, age and sex on shape variation (62–64).

Facial growth and ontogeny were examined through multivariate regressions of shape on facial size and age (65). Facial size was quantified as centroid size, defined as the square root of summed squared distances from landmarks to the centroid (65). When size explained a statistically significant percentage of shape variation, Procrustes distances were recalculated using the regression residuals of the multivariate regression to remove the effect of allometry. Additionally, a Procrustes ANOVA assessed the interaction between age and diagnosis for each syndrome. All morphometric and statistical analyses were performed using MorphoJ (66) and geomorph (62,63).

## RESULTS

### Distinct 3D morphological patterns in DS, MS and NS

The PCA revealed separation along PC1 between controls and individuals with all syndromes (DS, MS, NS), except for NF1 (Figure 2). In DS (Figure 2A), facial changes included a wider, flatter face; oblique palpebral fissures; hypertelorism; a depressed nasal bridge; and a broad nose with widely spaced alae (Figure 2A). Mandibular prognathism and thickened upper lips were also observed, with the latter emerging as a distinct characteristic in this population. Procrustes distance analysis confirmed significant shape differences between patients and controls (Procrustes distances= 0.0801; p <.0001).

**Figure 2.**
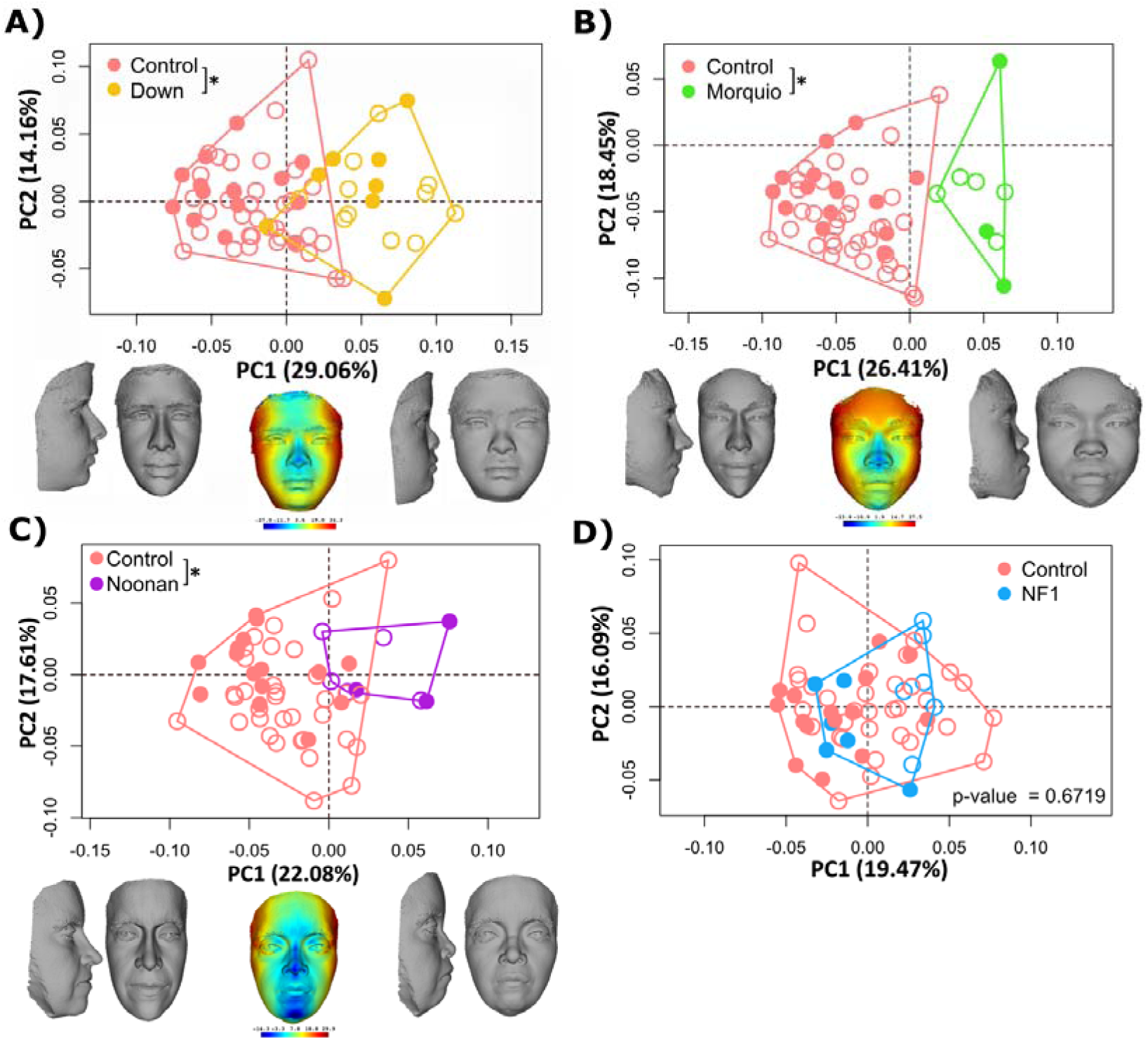
Principal Component Analyses (PCA) results comparing controls with DS (A), MS (B), NS (C), and NF1 (D). Female individuals indicated with empty symbols. Morphings show frontal and lateral facial shape changes along PC1. When facial shape differences between control and patients were significant (*) p < .001. Heatmaps indicate regions of greatest variation. The faces in the figure are morphings which do not belong to any real individual and are created from 3D landmark coordinates.

MS showed the most pronounced dysmorphology, with no overlap between patients and controls (Figure 2B). It also displayed the largest Procrustes distance between controls and patients (0.0943; p <.0001). Features included facial retrusion, increased interorbital distance, nasal bridge, a protruding forehead, and a thickened upper lip (Figure 2B).

NS presented milder differences, as indicated by a partial group overlap in PCA (Figure 2C) and the smallest Procrustes distance among affected groups (0.0683; p <.0001). NS was associated with a longer, narrower face, mandibular retrognathism, and thick lips. Neither mandibular retrognathism nor thick lips had been previously described in other populations.

NF1 was the only syndrome that showed no significant shape differences. PCA revealed complete overlap between patients and controls (Figure 2D), and Procrustes distances were not statistically significant (0.0212; p=0.6719).

### Continuous spectrum of facial dysmorphology

Analysis of the pooled sample (Figure 3) showed that the first two PCs accounted for 42.24% of total facial shape variation.

**Figure 3.**
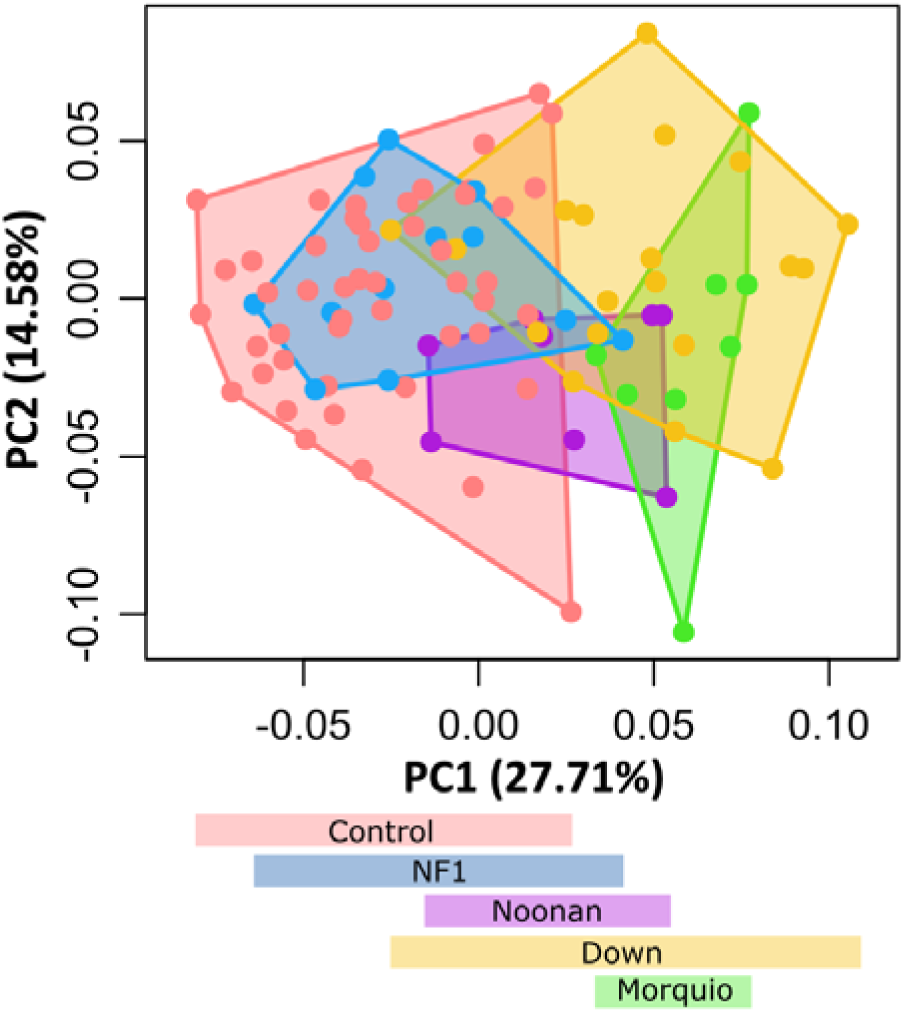
Principal Component Analysis (PCA) of pooled sample showing continuous variation in facial morphology across groups.

Although substantial overlap among groups was evident, positioning along PC1 reflected a gradient of dysmorphology. Control individuals clustered at the negative extreme, with NF1 cases largely overlapping and suggesting minimal dysmorphology associated with this rare disorder. NS individuals occupied an intermediate position, partially overlapping with both controls and NF1. In contrast, DS and MS cases were shifted toward the positive extreme, with DS showing the widest dispersion along PC1 and MS was the only syndrome that completely separated from controls (Figure 3).

### Factors modulating facial morphology

First, we analyzed separately the effect of facial size. The multivariate regression of centroid size on facial shape confirmed a significant allometric effect (p <.0001), with facial size explaining 15.4% of shape variation. Procrustes distances between groups recalculated from the regression residuals confirmed that after removing the effect of size, we still detected significant facial shape differences between controls and individuals with DS, MS, and NS (Table 2).

**Table 2:**
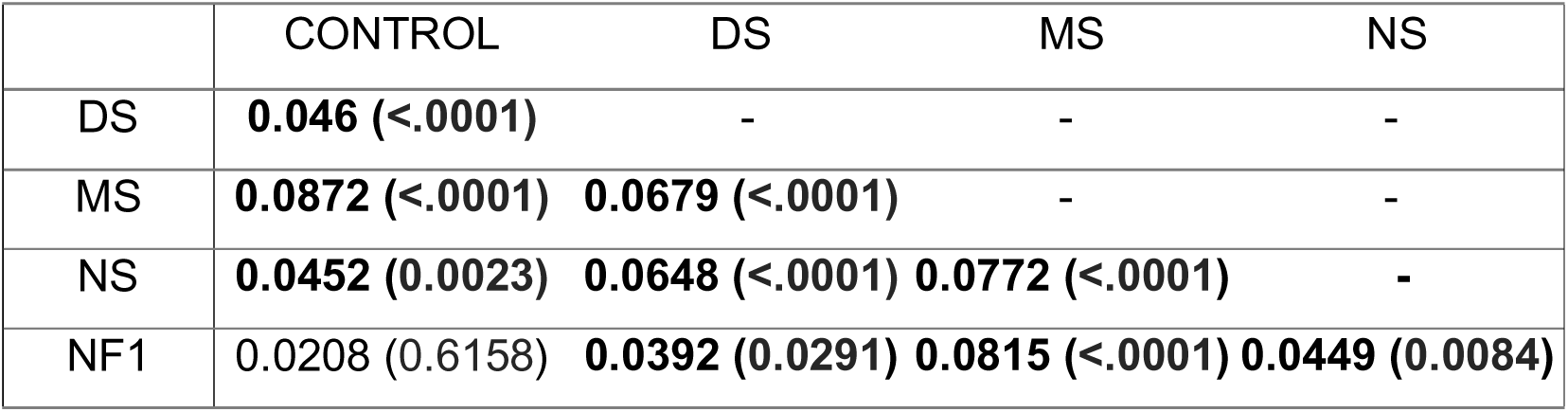
Procrustes distances and associated p-values based on regression residuals of facial shape on centroid size among diagnoses. Significant facial differences between groups are highlighted in bold.

|  | CONTROL | DS | MS | NS |
| --- | --- | --- | --- | --- |
| DS | **0.046 (<.0001)** | - | - | - |
| MS | **0.0872 (<.0001)** | **0.0679 (<.0001)** | - | - |
| NS | **0.0452 (0.0023)** | **0.0648 (<.0001)** | **0.0772 (<.0001)** | - |
| NF1 | 0.0208 (0.6158) | **0.0392 (0.0291)** | **0.0815 (<.0001)** | **0.0449 (0.0084)** |

Second, we performed a regression of facial shape on age to assess the effect of aging pooling all syndromes. Results indicated a significant positive correlation, with age explaining 9.7% of morphological variation (p <.0001). When analyzing each syndrome separately, we detected that in DS, NS and NF1 the trajectory was parallel to controls (Figure 4). The Procrustes ANOVA analyses showed that the interaction between age and diagnosis was not statistically significant (DS: p=0.639; NS: p=0.635; NF1: p=0.577), indicating a similar ontogenic development in these conditions as compared to controls. Only MS displayed a divergent trajectory and altered postnatal development (Figure 4B); however, the interaction factor did not reach significance (p=0.074), likely due to the small sample size (Additional file 2).

**Figure 4.**
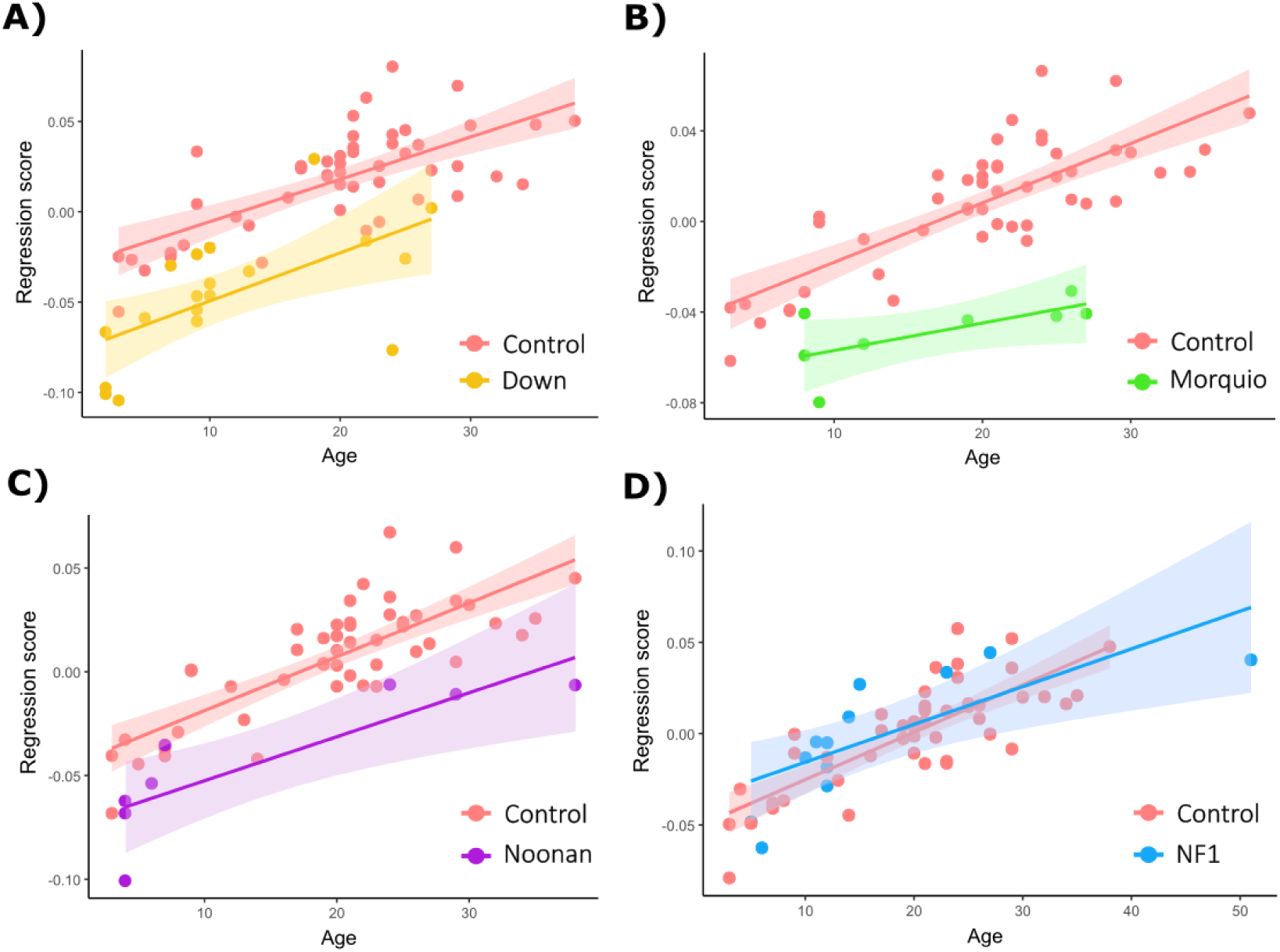
Ontogenetic trajectories of facial shape comparing controls with DS (A), MS (B), NS (C), and NF1 (D).

Finally, we assessed the simultaneous effect of all factors by performing a type II Procrustes ANOVA (Table 3), which estimates the portion of variance in facial shape that is attributable to each factor when the remaining covariates are adjusted for. All four factors (diagnosis, age, size and sex) showed a significant influence on facial shape, but diagnosis was the factor that explained most variance, 14% (Table 3). Facial size, age and sex only explained 1-2% of facial shape variation when diagnosis and the other factors had been accounted for.

**Table 3.**
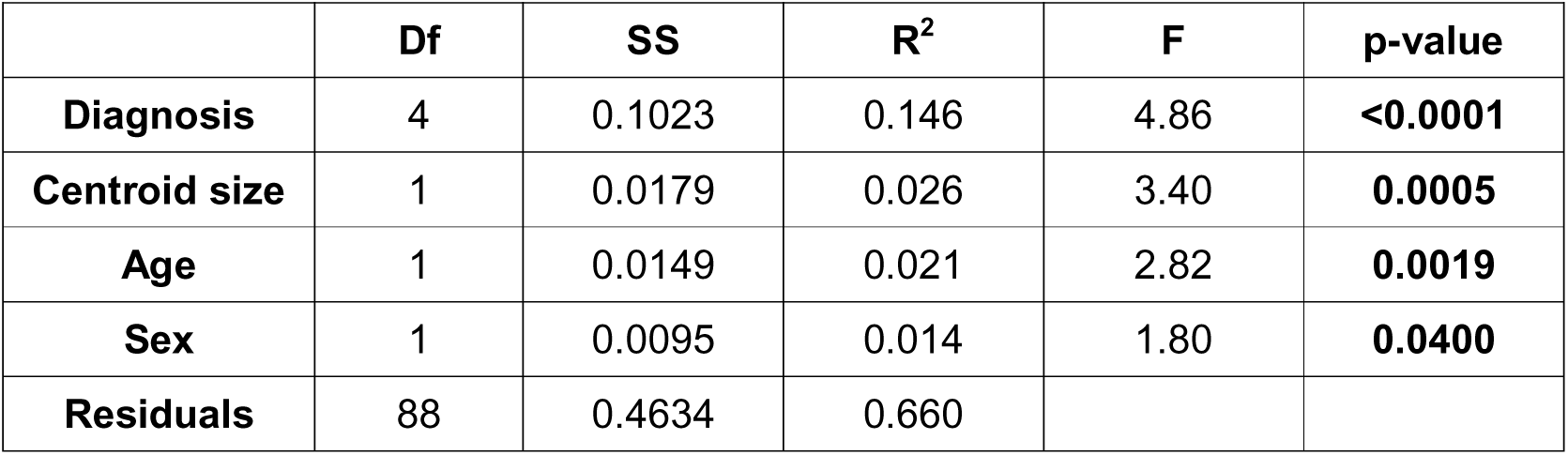
Procrustes ANOVA results showing the effects of diagnosis, centroid size, age, and sex on facial shape variation. Reported values include degrees of freedom (Df), sum of squares (SS), proportion of explained variance (R^2^), F-statistics, and permutation-based p-values.

| | Df | SS | $R^2$ | F | p-value |
| --- | --- | --- | --- | --- | --- |
| **Diagnosis** | 4 | 0.1023 | 0.146 | 4.86 | **<0.0001** |
| **Centroid size** | 1 | 0.0179 | 0.026 | 3.40 | **0.0005** |
| **Age** | 1 | 0.0149 | 0.021 | 2.82 | **0.0019** |
| **Sex** | 1 | 0.0095 | 0.014 | 1.80 | **0.0400** |
| **Residuals** | 88 | 0.4634 | 0.660 |  |  |

## DISCUSSION

This study extends previous analyses of facial dysmorphologies associated with DS, MS, NS, and NF1 in a Colombian population (7). The use of 3D geometric morphometric methods represents a significant advancement over qualitative or 2D quantitative approaches, providing a more precise and quantitative means of detecting facial shape differences while controlling for confounding factors such as facial size and age (11). Unlike 2D methods, 3D analyses incorporate depth, a critical shape component for accurately capturing syndrome-specific dysmorphologies and thus enhance the potential application of facial biomarkers for diagnosis and prognosis of genetic and rare disorders (12–14,21–27).

Our findings indicate that, although Colombian individuals share general dysmorphological patterns reported in other populations, they also display population-specific traits, reflecting the diverse genetic ancestry of this Latin-American population (7). In Cali, admixture with the indigenous Amerindian population began in the sixteenth century with the arrival of Spanish colonizers (67). In the eighteenth century, the establishment of large colonial settlements of African slaves for sugar cane exploitation significantly changed the population structure of Valle del Cauca (67). Currently, the Amerindian and African ancestry predominate in this Colombian region over European ancestry (68).

For DS, observed features largely align with reports from European-descent populations (36,38,39,41), but a thicker upper lip relative to the lower lip emerges as a distinct characteristic in this Latin American population. In MS, all traits described by Suárez-Guerrero et al. (2020) were present, including the same lip thickness difference noted in DS. Our findings for NS diverged from European-based descriptions (50), with Colombian individuals showing mandibular retrognathism and thick lips, which are features not consistently reported elsewhere. For NF1, no significant differences were detected, consistent with prior studies (54), indicating subtle, non-significant facial variation in this syndrome. Although previous research suggested shared dysmorphologies among RASopathies (46), our results support the differentiation of NS and NF1 based on facial phenotypes.

Pooling all syndromic groups revealed a continuous spectrum of facial variation: NS showed mild to moderate changes, DS moderate to severe, and MS the most severe dysmorphologies. These findings expand upon earlier 2D analyses (7), revealing additional 3D shape details and allometric effects. 3D analyses allowed for the identification of depth-related traits, such as mandibular prognathism or retrognathism, while enabling the visualization of syndrome-specific facial morphology through 3D morphs. The observed variation in explanatory power across variables reflects shared variance and interaction effects, emphasizing the need for multifactorial modeling.

Analysis of ontogenetic trajectories showed that NS, NF1 and DS followed similar growth patterns to controls. In contrast, MS exhibited deviations, suggesting altered postnatal facial development. This result is consistent with previous evidence describing that, in mucopolysaccharidosis syndromes, facial dysmorphologies of MS are cumulative and increase over lifetime due to progressive lysosomal accumulation of glycosaminoglycans (GAGs) (42). However, statistical significance was not reached when evaluating the interaction between age and diagnosis, likely due to the small sample size. Overall, these results underscore the need to specifically assess ontogeny to improve the prognosis and clinical management of individuals with genetic and rare disorders.

Research on rare conditions is often constrained by their low prevalence, making it crucial to expand both sample size and diversity (7,13). Broader representation across different ages, sexes, and populations from around the world would provide a more comprehensive understanding of phenotypic variability. Larger datasets would support more refined analyses of facial phenotypes, the identification of morphological differences based on facial distances, and the early recognition of dysmorphologies during childhood development.

Increasing diversity in biomedical research would not only enhance current diagnostic models but also improve the identification and mapping of genetic variants associated with anthropometric traits and diseases (69). Incorporating ancestral diversity into biomedical studies would enable the establishment of more precise and specific biological profiles for each syndrome, considering the impact of ancestry on facial phenotype (15). Moroever, integrating genetic data with phenotypic analyses could enable the identification of genotype-phenotype correlations, which are critical for a deeper understanding of molecular pathways and phenotypic subgroups associated with genetic disorders, improving genetic variant interpretation, ultimately supporting precise diagnosis and personalized care (70).

In the future, translating facial biomakers into clinical practice will require the development and validation of low-cost 3D facial shape acquisition methodologies (71). Such tools have the potential to considerably shorten the diagnostic odyssey for rare diseases, particularly in regions where access to genetic testing remains limited.

## CONCLUSION

Overall, our 3D morphometric analyses confirmed distinct diagnostic patterns in DS, MS and NS. This finding highlights the potential of facial shape analysis to aid the diagnosis of these genetic conditions, and the importance to incorporate ontogenetic and interpopulation variation to optimize their clinical applicability across diverse human populations. In regions with limited access to genetic testing, these morphometric approaches could offer a non-invasive, affordable, and accessible alternative to support early diagnosis and clinical decision-making.

## Data Availability

Raw phenotype data from the Colombian population cannot be made available due to restrictions imposed by the ethics approval.

## LIST OF ABBREVIATIONS

3D: Three dimensional
DS: Down syndrome
MS: Morquio syndrome
NS: Noonan syndrome
NF1: Neurofibromatosis type 1
2D: Two dimensional
SHH: Sonic Hedgehog
FGF: Fibroblast Growth Factor
BMP: Bone Morphogenetic Protein
TGF β: Transforming Growth Factor β
GPA: Generalized Procrustes Analysis
PCA: Principal Components Analysis
PCs: Principal Components

## DECLARATIONS

### Ethics approval and consent to participate

The study was approved by the Ethics Committee for Human Research at Universidad Icesi (approval record no. 309), the Bioethics Committee of the Universitat de Barcelona (IRB03099-CER032514) and adhered to national guidelines and protocols, following the standards for medical research in humans recommended by the Declaration of Helsinki. All participants provided informed consent before enrollment. For minors, consent was obtained from parents or legal guardians for the acquisition of facial photographs and associated clinical data.

### Consent for publication

The faces used in this manuscript for illustrative purposes do not belong to any real individual and correspond to virtual models created from random facial meshes morphed by 3D landmark coordinates defining axes of morphological variation. Therefore, no consent for publication is required.

### Availability of data and materials

Raw image data cannot be made available due to restrictions imposed by ethics approval. Landmark data supporting the findings of this study are available from the corresponding author upon reasonable request.

### Competing interests

The authors declare that they have no competing interests.

### Funding

This study was funded by Proyecto COL0012168-1097 Interfacultades-ICESI, Beca predoctoral Fundación Álvaro Entrecanales-Lejeune to LME-Q, the Agencia Española de Investigación (projects PID2023-147001OB-C21/C22 funded by MICIU/AEI/10.13039/501100011033 and by ERDF/EU), Programa de Contractació d’Investigadors Predoctorals en Formació de la Universitat de Barcelona (PREDOCS-UB 2024), Wenner Gren Foundation for Anthropological Research (Gr. 10657), Agència de Gestió d’Ajuts Universitaris i de Recerca (AGAUR) of the Generalitat de Catalunya (2021 SGR00706 and 2021 SGR01396), and Biological Anthropological Master UB-UAB.

### Author’s contributions

MAM, MP, EE, AC, HP and NMA conceived the project. LMEQ, EC, EG, ORR, MRT and HP recruited participants, performed 3D facial scanning and diagnosed cases. MAM, MP, AHL, XS, AC and NMA performed the morphometric analyses and interpreted results. MAM, MP, and NMA prepared the manuscript and figures. All authors critically reviewed the manuscript.

## Acknowledgements

We are grateful for the voluntary collaboration of all participants, including children and their families. We are thankful to Colegio Ecológico Scout and Universidad Icesi for granting us permission to organize the photographic sessions in Colombia.

## Notes

### Competing Interest Statement

The authors have declared no competing interest.

### Funding Statement

This study was funded by Proyecto COL0012168-1097 Interfacultades-ICESI, Beca predoctoral Fundacion
Alvaro Entrecanales-Lejeune to LME-Q, Joan Oro grant (2024 FI-3 00160) from the Recerca
i Universitats Departament (DRU) of the Generalitat de Catalunya and
the European Social Fund to AH-L, Wenner Gren Foundation for Anthropological Research (Gr. 10657), Agencia de Gestio Ajuts Universitaris i de Recerca (AGAUR) of the
Generalitat de Catalunya (2021 SGR00706 and 2021 SGR0139), and Biological Anthropological Master UB-UAB.

### Author Declarations

The study was approved by the Ethics Committee for Human Research at Universidad Icesi with approval record no. 309, and complied with the national guidelines and protocols.

### Summary of Updates

In the revised version of the manuscript, we have included new analyses, such as Procrustes ANOVA to quantify the amount of facial shape variation attributable to modulating factors and associated figures. The text has also been been reviewed and proof-read. The author list has been updated to reflect the changes.

