## Additional file 2 for "3D phenotyping in a Colombian population reveals unique population and ontogenic facial patterns in genetic and rare disorders"

**Additional file 2.** Procrustes ANOVA results showing the interaction between age and diagnosis. Reported values include degrees of freedom (Df), sum of squares (SS), proportion of explained variance (R^2^), F-statistics, and permutation-based p-values.

|  | **Df** | **SS** | **R^2^** | **F** | **p-value** |
| --- | --- | --- | --- | --- | --- |
| **DS** | 1 | 0.0045 | 0.0093 | 0.8141 | 0.6387 |
| **MS** | 1 | 0.0088 | 0.0238 | 1.4411 | 0.0738 |
| **NS** | 1 | 0.0040 | 0.0122 | 0.8095 | 0.6351 |
| **NF1** | 1 | 0.0044 | 0.0133 | 0.8613 | 0.5772 |
