## Additional file 1 for "3D phenotyping in a Colombian population reveals unique population and ontogenic facial patterns in genetic and rare disorders"

**Additional file 1.** Anatomic definition of the facial landmarks collected from the 3D models shown in Figure 1.

| **REFERENCE POINT** | **ANATOMICAL POSITION** |
| --- | --- |
| 1 | **Glabellla:** midpoint between the eyebrows |
| 2 | **Sellion:** deepest point of the nasal root |
| 3 | **Pronasale:** most anterior point of the tip of the nose |
| 4 | **Subnasale:** point where the nasal septum meets the nasal groove |
| 5 | **Labiale Superius:** midpoint of the superior lip suture |
| 6 | **Labial Inferius:** midpoint of the inferior lip suture |
| 7 | **Gnathion:** Most inferior point of the chin |
| 8 | **Endocanthion R:** inner lateral commissure point of the right eye |
| 9 | **Endocanthion L:** inner lateral commissure point of the left eye |
| 10 | **Palpebral inferior R:** most inferior medial point of the right lower eyelid |
| 11 | **Exocanthion R:** outer lateral commissure point of the right eye |
| 12 | **Palpebral inferior L:** most inferior medial point of the left lower eyelid |
| 13 | **Exocanthion L:** outer lateral commissure point of the left eye |
| 14 | **Alare R:** most lateral point of the right nasal wing |
| 15 | **Subalare R:** facial insertion of the right alar base |
| 16 | **Subalare L:** facial insertion of the left alar base |
| 17 | **Alare L:** most lateral point of the left nasal wing |
| 18 | **Chelion R:** point located at the right labial commissure |
| 19 | **Crista philtra R:** intersection of the vermilion line and the raised edge of the right nasolabial groove |
| 20 | **Crista philtra L:** intersection of the vermilion line and the raised edge of the left nasolabial groove |
| 21 | **Chelion L:** point located at the left labial commissure |
